# Machine learning prediction for early-stage melanoma outcomes: recurrence-free survival, disease-specific survival, and overall survival

**DOI:** 10.1101/2025.05.28.25328519

**Authors:** Guihong Wan, Hannah Rashdan, Olivia M. Burke, Sara Khattab, Nga Nguyen, Bonnie W. Leung, Emma Beagles, Crystal T. Chang, Kun-Hsing Yu, Mia S. DeSimone, Yevgeniy R. Semenov

**Author notes:** **Corresponding author:** Yevgeniy R. Semenov, MD, MA, Department of Dermatology, Massachusetts General Hospital, Harvard Medical School, 40 Blossom Street, Bartlett Hall 6R, Room 626, Boston, MA 02114. Designates co-first authors (Guihong Wan, Hannah Rashdan). Designates co-senior authors (Mia S. DeSimone, Yevgeniy R. Semenov).

## Abstract

This study compared machine-learning models for predicting recurrence-free survival (RFS), disease-specific survival (DSS), and overall survival (OS) using clinicopathologic data from 1,621 stage I/II primary cutaneous melanoma patients. Our time-to-event models achieved concordance indices of 0.829 for RFS, 0.812 for DSS, and 0.778 for OS. Tumor thickness and mitotic rate were the most important predictors for RFS. Charlson comorbidity score and insurance type were critical for DSS and OS.

---

Melanoma is a highly aggressive cancer with a substantial risk of recurrence. Most patients with melanoma recurrence had the disease that was initially early-stage (stage I/II) at diagnosis.^1^ Tumor thickness, clinical stage, mitotic rate, age at diagnosis, melanoma location, and ulceration status have been associated with melanoma recurrence.^2,3^ However, widely adopted, comprehensive risk stratification approaches for early-stage melanoma patients remain lacking, with the current reliance on AJCC staging alone offering limited predictive accuracy for identifying patients at the highest risk of recurrence.^4^

The recent approval by the US Food and Drug Administration of adjuvant immune checkpoint inhibitor (ICI) therapy for managing stage IIB and IIC melanoma underscores the need for risk stratification approaches to identify early-stage melanoma patients with more aggressive disease for whom the potential benefits of immunotherapy may outweigh risk of toxicities associated with this therapeutic class. The extension of ICI therapy to this population was based on clinical trial results demonstrating improved recurrence-free survival (RFS) among patients treated with adjuvant ICIs compared to those who only underwent surgical excision.^5^ While RFS is an important endpoint, it has limitations, particularly in a setting where most recurrences are localized and are, therefore, not expected to impact mortality directly.^2^ Considering that a major goal of cancer treatment is to extend survival, disease-specific survival (DSS) and overall survival (OS) are considered gold standard endpoints in oncology,^6^ but typically require trials with larger patient populations and longer follow-up. Thus, it is crucial to examine the extent to which early-stage melanoma risk stratification and management using RFS as an endpoint can be reliably extended to predict DSS and OS.

In this study, we aimed to address these gaps by comparing machine-learning models for predicting RFS, DSS, and OS, and investigating whether models trained on RFS data can also reliably estimate DSS and OS in this population.

## Patient characteristics

This study identified 1,621 patients diagnosed with stage I/II primary cutaneous melanoma at Massachusetts General Hospital (MGH) and Brigham and Women’s Hospital/Dana-Farber Cancer Institute (BWH/DFCI) between 2000 and 2020. We extracted 31 clinicopathologic features from patients’ electronic health records. The characteristics of the study population are presented in **Table 1**, with a complete list of clinicopathologic features provided in **Supplementary Table 1**. In the MGH cohort, 217 (19.7%) patients experienced disease recurrence, 106 (9.6%) died from melanoma, and a total of 382 (34.7%) were dead at the last follow-up. In the BWH/DFCI cohort, 91 (17.5%) patients experienced disease recurrence, 33 (6.3%) died from melanoma, and a total of 105 (20.2%) were dead at the last follow-up. The cumulative incidence of melanoma recurrence, melanoma-specific mortality, and overall mortality are presented in **Figure 1**. The melanoma-specific mortality rate was higher among patients with distant recurrence compared to patients with locoregional recurrence (57.9% vs 32.7%, P-value<0.001). Compared to patients with non-recurrent melanoma, patients with recurrent melanoma had a higher risk of mortality (HR: 2.76; P-value<0.001) (**Supplementary Figure 1).**

**Table 1.**
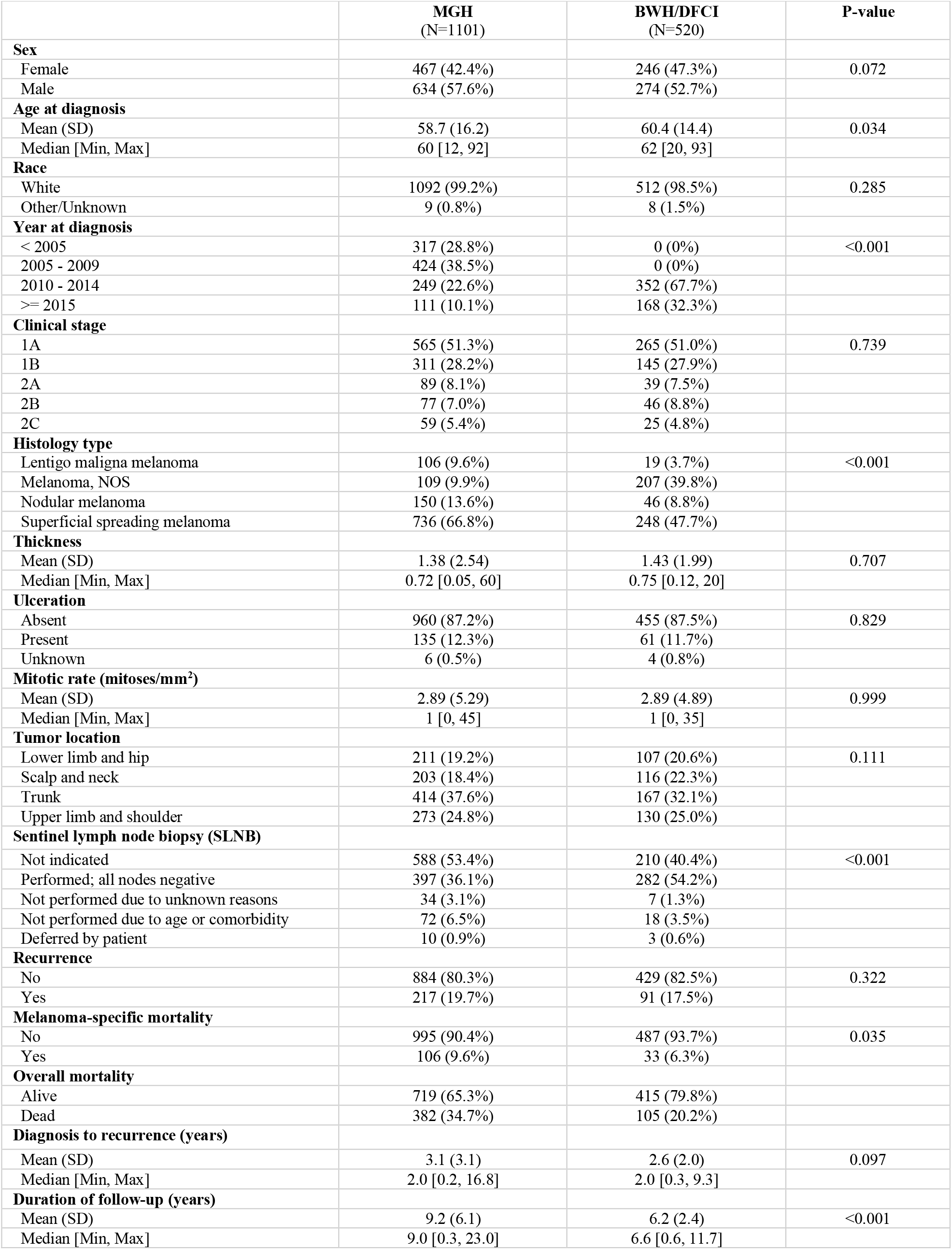
Characteristics of the study population

|  | **MGH**<br>(N=1101) | **BWH/DFCI**<br>(N=520) | **P-value** |
| --- | --- | --- | --- |
| **Sex** |  |  |  |
| Female | 467 (42.4%) | 246 (47.3%) | 0.072 |
| Male | 634 (57.6%) | 274 (52.7%) |  |
| **Age at diagnosis** |  |  |  |
| Mean (SD) | 58.7 (16.2) | 60.4 (14.4) | 0.034 |
| Median [Min, Max] | 60 [12, 92] | 62 [20, 93] |  |
| **Race** |  |  |  |
| White | 1092 (99.2%) | 512 (98.5%) | 0.285 |
| Other/Unknown | 9 (0.8%) | 8 (1.5%) |  |
| **Year at diagnosis** |  |  |  |
| < 2005 | 317 (28.8%) | 0 (0%) | <0.001 |
| 2005 - 2009 | 424 (38.5%) | 0 (0%) |  |
| 2010 - 2014 | 249 (22.6%) | 352 (67.7%) |  |
| >= 2015 | 111 (10.1%) | 168 (32.3%) |  |
| **Clinical stage** |  |  |  |
| 1A | 565 (51.3%) | 265 (51.0%) | 0.739 |
| 1B | 311 (28.2%) | 145 (27.9%) |  |
| 2A | 89 (8.1%) | 39 (7.5%) |  |
| 2B | 77 (7.0%) | 46 (8.8%) |  |
| 2C | 59 (5.4%) | 25 (4.8%) |  |
| **Histology type** |  |  |  |
| Lentigo maligna melanoma | 106 (9.6%) | 19 (3.7%) | <0.001 |
| Melanoma, NOS | 109 (9.9%) | 207 (39.8%) |  |
| Nodular melanoma | 150 (13.6%) | 46 (8.8%) |  |
| Superficial spreading melanoma | 736 (66.8%) | 248 (47.7%) |  |
| **Thickness** |  |  |  |
| Mean (SD) | 1.38 (2.54) | 1.43 (1.99) | 0.707 |
| Median [Min, Max] | 0.72 [0.05, 60] | 0.75 [0.12, 20] |  |
| **Ulceration** |  |  |  |
| Absent | 960 (87.2%) | 455 (87.5%) | 0.829 |
| Present | 135 (12.3%) | 61 (11.7%) |  |
| Unknown | 6 (0.5%) | 4 (0.8%) |  |
| **Mitotic rate (mitoses/mm<sup>2</sup>)** |  |  |  |
| Mean (SD) | 2.89 (5.29) | 2.89 (4.89) | 0.999 |
| Median [Min, Max] | 1 [0, 45] | 1 [0, 35] |  |
| **Tumor location** |  |  |  |
| Lower limb and hip | 211 (19.2%) | 107 (20.6%) | 0.111 |
| Scalp and neck | 203 (18.4%) | 116 (22.3%) |  |
| Trunk | 414 (37.6%) | 167 (32.1%) |  |
| Upper limb and shoulder | 273 (24.8%) | 130 (25.0%) |  |
| **Sentinel lymph node biopsy (SLNB)** |  |  |  |
| Not indicated | 588 (53.4%) | 210 (40.4%) | <0.001 |
| Performed; all nodes negative | 397 (36.1%) | 282 (54.2%) |  |
| Not performed due to unknown reasons | 34 (3.1%) | 7 (1.3%) |  |
| Not performed due to age or comorbidity | 72 (6.5%) | 18 (3.5%) |  |
| Deferred by patient | 10 (0.9%) | 3 (0.6%) |  |
| **Recurrence** |  |  |  |
| No | 884 (80.3%) | 429 (82.5%) | 0.322 |
| Yes | 217 (19.7%) | 91 (17.5%) |  |
| **Melanoma-specific mortality** |  |  |  |
| No | 995 (90.4%) | 487 (93.7%) | 0.035 |
| Yes | 106 (9.6%) | 33 (6.3%) |  |
| **Overall mortality** |  |  |  |
| Alive | 719 (65.3%) | 415 (79.8%) |  |
| Dead | 382 (34.7%) | 105 (20.2%) |  |
| **Diagnosis to recurrence (years)** |  |  |  |
| Mean (SD) | 3.1 (3.1) | 2.6 (2.0) | 0.097 |
| Median [Min, Max] | 2.0 [0.2, 16.8] | 2.0 [0.3, 9.3] |  |
| **Duration of follow-up (years)** |  |  |  |
| Mean (SD) | 9.2 (6.1) | 6.2 (2.4) | <0.001 |
| Median [Min, Max] | 9.0 [0.3, 23.0] | 6.6 [0.6, 11.7] |  |

**Figure 1.**
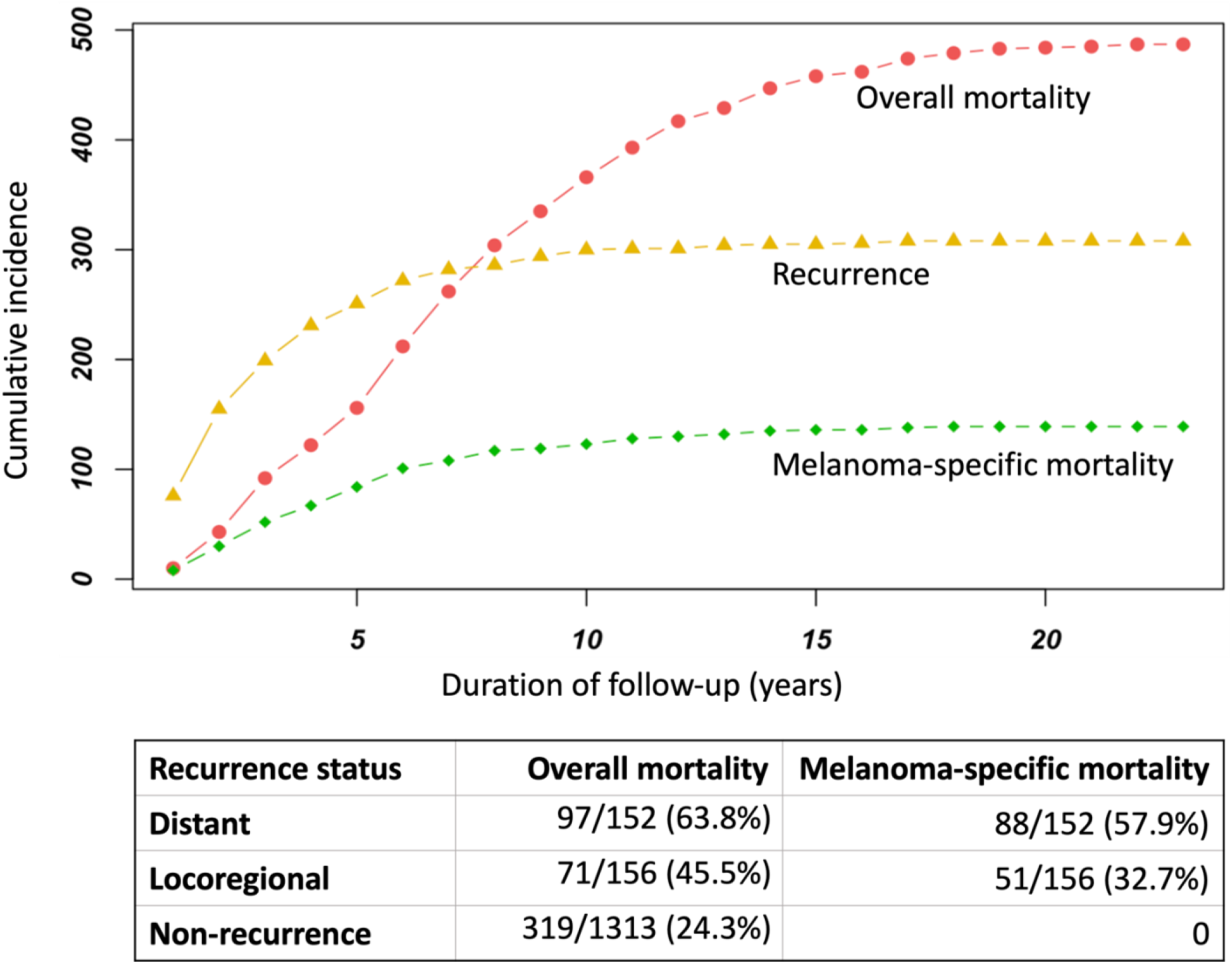
Cumulative incidence of melanoma recurrence, melanoma-specific mortality, and overall mortality **Legend:** The cumulative incidence pattern of melanoma-specific mortality over time is similar to that of melanoma recurrence, while the cumulative incidence pattern of overall mortality is different compared to that of melanoma recurrence. Compared to locoregional recurrence, the Hazard Ratios of distant recurrence on overall mortality and melanoma-specific mortality are 2.64 (95% CI: 2.03-3.44; p<0.001) and 8.83 (95% CI: 5.87-13.3; p<0.001), respectively.

## Machine-learning model performance

To predict RFS, DSS, and OS, we used two time-to-event machine-learning algorithms, Gradient Boosting Survival (GBS)^10^ and Random Survival Forest (RSF),^11^ the best performers in our previous studies.^2,7^ Models were evaluated by concordance index^12^ using (1) five-fold cross-validation of the MGH cohort (internal validation); (2) the MGH cohort for model training and the BWH/DFCI cohort for testing the models independently (external validation). We also examined the transferability of the model trained for recurrence prediction on the MGH cohort to estimate the risk of melanoma-specific and overall mortality on the BWH/DFCI cohort (cross-transfer validation).

**Table 2** presents model performance for predicting melanoma outcomes. In the RFS prediction, the best performance was 0.877 (95% CI: 0.871-0.882) and 0.829 (95% CI: 0.827-0.831) in the internal and external validations, respectively. The best performance in the DSS prediction was 0.875 (95% CI: 0.862-0.888) and 0.812 (95% CI: 0.810-0.814) in the internal and external validations, respectively. In the OS prediction, the best performance was 0.826 (95% CI: 0.821-0.831) and 0.778 (95% CI: 0.777-0.781) in the internal and external validations, respectively. In the cross-transfer validation, the model performance deteriorated significantly for DSS (0.789; 95% CI: 0.786-0.792) and, to a greater extent, OS (0.631; 95% CI: 0628-0.634).

**Table 2.** Prediction of melanoma outcomes

|  | Prediction | Internal Validation<br>(mean and 95% CI) |  | External Validation<br>(mean and 95% CI) |  |
| --- | --- | --- | --- | --- | --- |
|  |  | RSF | GBS | RSF | GBS |
| Reference<sup>1</sup> | RFS | 0.870<br>0.863-0.876 | 0.877<br>0.871-0.882 | 0.804<br>0.802-0.806 | **0.829**<br>**0.827-0.831** |
| Individual<sup>2</sup> | DSS | 0.873<br>0.860-0.886 | 0.875<br>0.862-0.888 | 0.783<br>0.777-0.789 | **0.812**<br>**0.810-0.814** |
|  | OS | 0.826<br>0.823-0.828 | 0.826<br>0.821-0.831 | 0.776<br>0.774-0.779 | **0.778**<br>**0.777-0.781** |
| Transfer<sup>3</sup> | DSS | N/A | N/A | 0.749<br>0.745-0.752 | **0.789**<br>**0.786-0.792** |
|  | OS | N/A | N/A | 0.614<br>0.611-0.619 | **0.631**<br>**0.628-0.634** |

<sup>1</sup>Models were trained and evaluated with RFS as the outcome.
<sup>2</sup>Models were trained and evaluated with DSS and OS as the outcome, respectively.
<sup>3</sup>The model was trained with RFS as the outcome. The same model was evaluated with DSS and OS as the outcome.
N/A: not applicable

We also investigated the critical features in the predictions by conducting permutation importance (**Supplementary Figures 2 and 3**). In both GBS and RFS models, thickness and mitotic rate were the most important features in the RFS prediction, while Charlson comorbidity score (CCS) is dominantly predictive for OS. In the DSS prediction, thickness and mitotic rate were ranked higher compared to the OS prediction in both GBS and RFS models.

## Discussion

This large-scale retrospective cohort study compares the predictive performance of machine-learning models for RFS, DSS, and OS in early-stage melanoma population. The models achieved similar performance for RFS (concordance index: 0.829), DSS (concordance index: 0.812), and OS (concordance index: 0.778) in the external validation. These results demonstrate that machine-learning models using a fixed set of features can independently predict outcomes of patients with early-stage melanoma.

The performance significantly declined when using the same model trained for predicting recurrence (concordance index: 0.829) to estimate the risk of melanoma-specific mortality and overall mortality (concordance index: 0.789 and 0.631, respectively). This raises concerns regarding the inherent assumption that the same factors that contribute to recurrence also lead to increased melanoma-specific mortality and overall mortality. As such, risk stratification utilizing disease recurrence as a prognostic endpoint can be extended to an extent to predict melanoma-specific mortality but not overall mortality in this population. Considering that recurrence-free survival is the outcome used in many clinical studies guiding oncologic algorithms, including in the most recent approval of ICIs for stage IIB/IIC melanoma,^5^ clinicians must practice caution when drawing conclusions regarding melanoma-specific mortality and overall mortality using the same features as for recurrence-free survival.

Furthermore, we found tumor thickness and mitotic rate were primary features in recurrence prediction, whereas CCS was the most notable feature in overall mortality prediction. Despite near-universal reliance of clinical algorithms on Breslow thickness and stage for prognostication,^1^ our results demonstrate limitations of predicting melanoma recurrence and mortality with current tools. Further risk stratification evaluating additional distinguishing features associated with melanoma recurrence and mortality is warranted.

## Methods

This study builds on recent studies from our group, which developed time-to-event machine-learning approaches for predicting disease recurrence in early-stage melanoma.^2,7^ We identified a retrospective cohort of 1,621 patients with stage I/II primary cutaneous melanoma diagnosed between January 2000 and February 2020, among which 1,101 were from Massachusetts General Hospital (MGH), and 520 were from Brigham and Women’s Hospital/Dana-Farber Cancer Institute (BWH/DFCI). We extracted 31 clinicopathologic features, including 10 clinical features and 21 tumor characteristics (**Supplementary Table 1**), and patient outcomes, including recurrence (status and date of first recurrence), melanoma-specific mortality, and overall mortality (status, death reason, and death date or last follow-up). Inclusion criteria and data extraction were consistent with our previous studies,^2,4,7^ except that the diagnosis date of the first melanoma was retained for patients with multiple stage I/II melanomas.

To compare groups, we used Pearson’s chi-squared test and Student’s t-test for categorical and continuous variables, respectively. Kaplan-Meier curves examined differences in overall survival stratified by melanoma recurrence type. To account for guarantee-time bias, Hazard Ratios (HRs) of melanoma recurrence on survival were computed using time-varying Cox proportional hazards models,^8,9^ adjusted for baseline demographics and tumor characteristics.

To predict RFS, DSS, and OS, we used two time-to-event machine-learning algorithms, GradientBoostingSurvivalAnalysis (GBS)^10^ and RandomSurvivalForest (RSF),^11^ the best performers in our previous studies.^2,7^ Models were evaluated by concordance index^12^ using (1) five-fold cross-validation of the MGH cohort (internal); (2) the MGH cohort for model training and the BWH/DFCI cohort for independent validation (external). We also examined the transferability of the model trained on the MGH cohort for recurrence prediction to estimate the risk of melanoma-specific and overall mortality on the BWH/DFCI cohort (cross-transfer validation). Each experiment was repeated 20 times. Mean and 95% confidence interval (CI) were reported. We further ranked features in the prediction by conducting permutation importance with 20 repeats. Experiments were implemented using the Python module scikit-survival 0.18.0.^13^

## DATA AVAILABILITY

All relevant data are available from the corresponding author: Yevgeniy R. Semenov. All summary data supporting the findings of this study are available within the article and/or its supplementary materials. The patient data generated for this study can only be shared per specific institutional review board (IRB) requirements. Upon a request to the corresponding author, a data-sharing agreement can be initiated following institution-specific guidelines.

## ACKNOWLEDGMENTS

GW is supported by the National Cancer Institute of the National Institutes of Health under Award Number K99CA286966. YRS is supported in part by the Department of Defense under Award Number W81XWH2110819, the National Institute of Arthritis and Musculoskeletal and Skin Diseases of the National Institutes of Health under Award Number K23AR080791, and the Melanoma Research Alliance Young Investigator Award.

## AUTHOR CONTRIBUTIONS

GW, HR, MSD, and YRS had full access to all the data in the study and took responsibility for the integrity of the data and the accuracy of the data analysis. Study concept and design: GW, HR, MSD, and YRS. Data acquisition: HR, OMB, SK, NN, GW, BWL, EB, CTC, BY, MSD, and YRS. Data analysis and interpretation: GW, HR, MSD, and YRS. Drafting of the manuscript: GW, HR, SK, MSD, and YRS. Administrative, technical, or material support: YRS. The study was conducted under the supervision of MSD and YRS. All authors reviewed, edited, and approved the manuscript.

---

**Supplementary Table 1.**
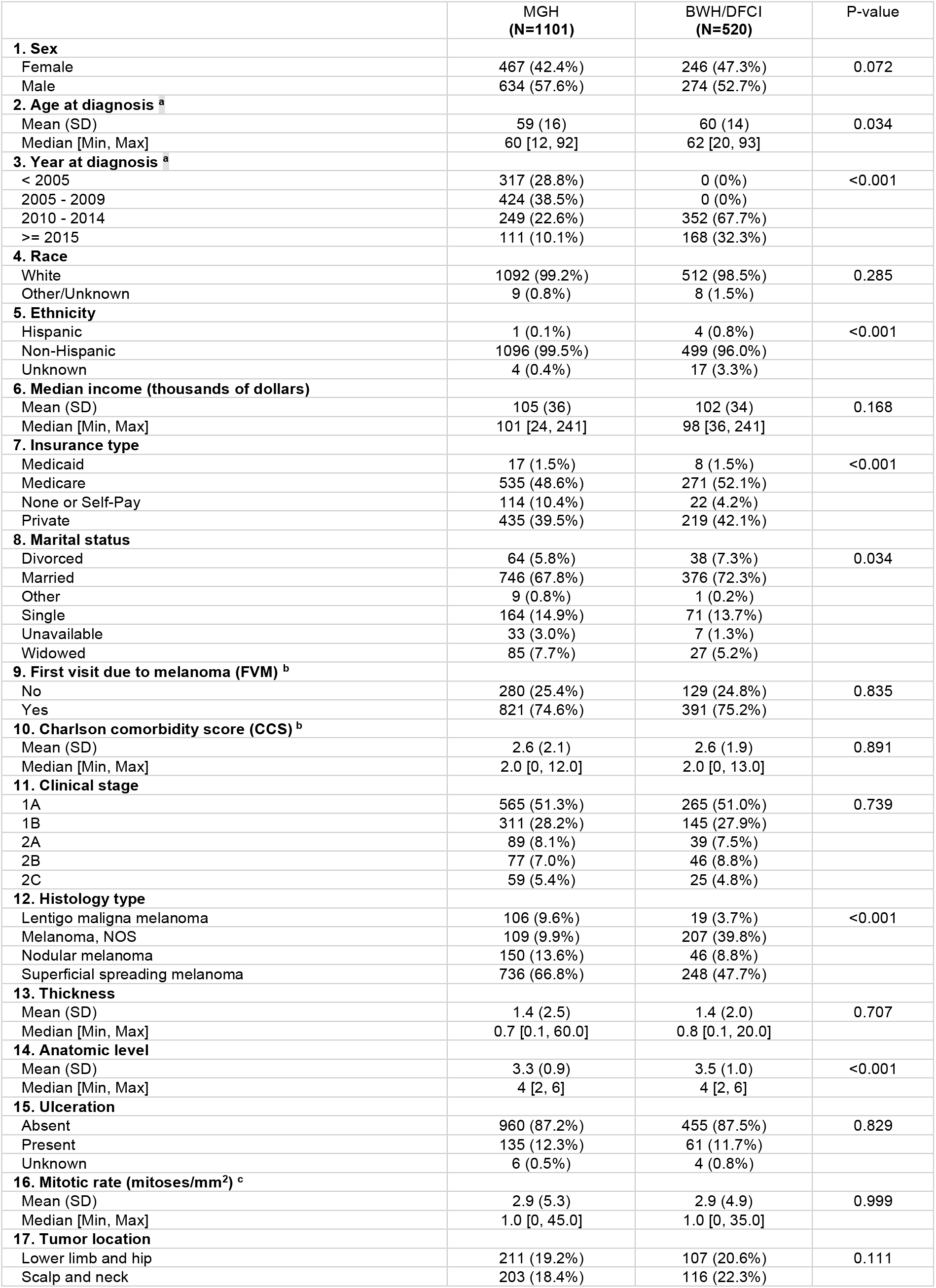
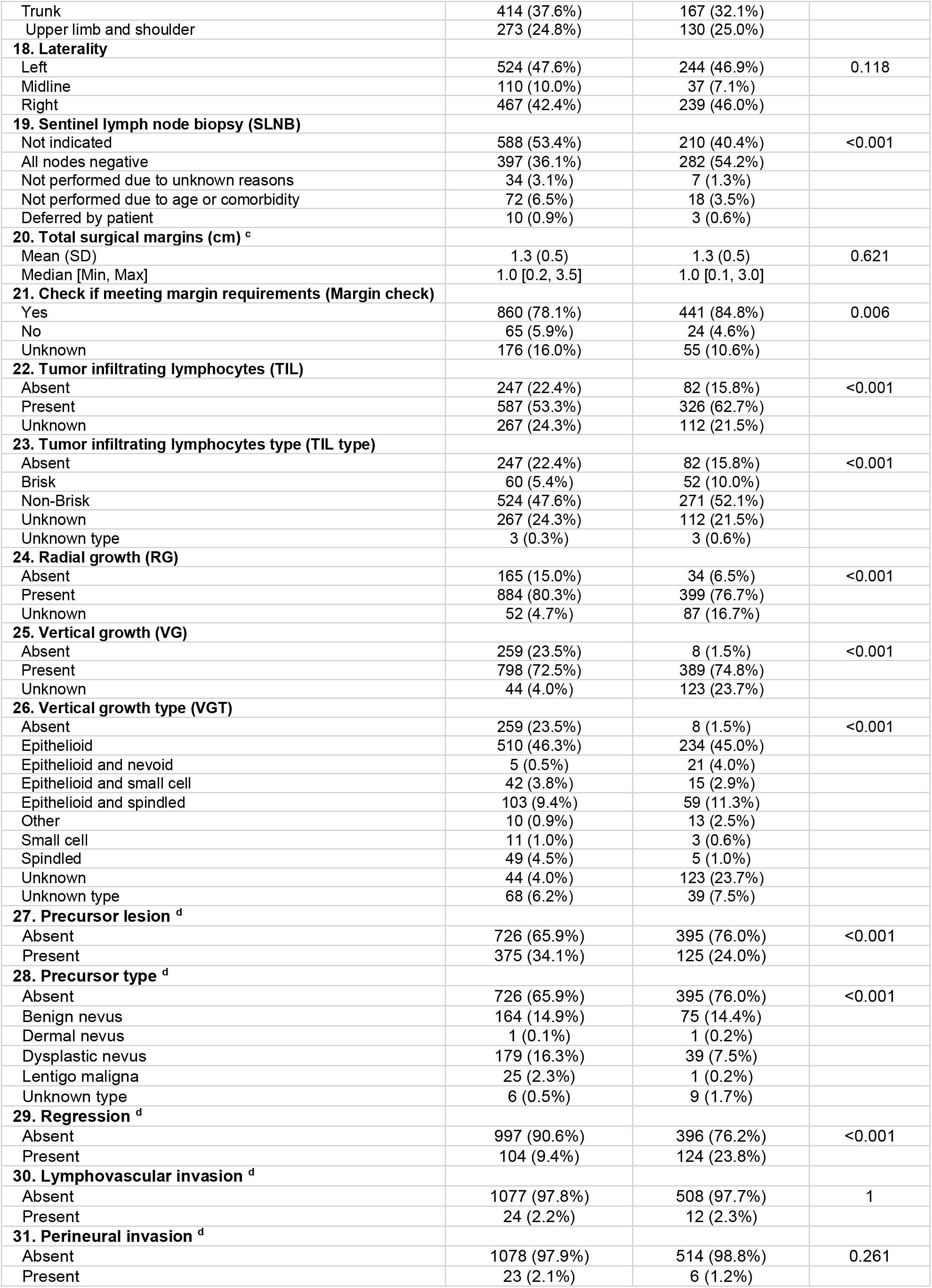
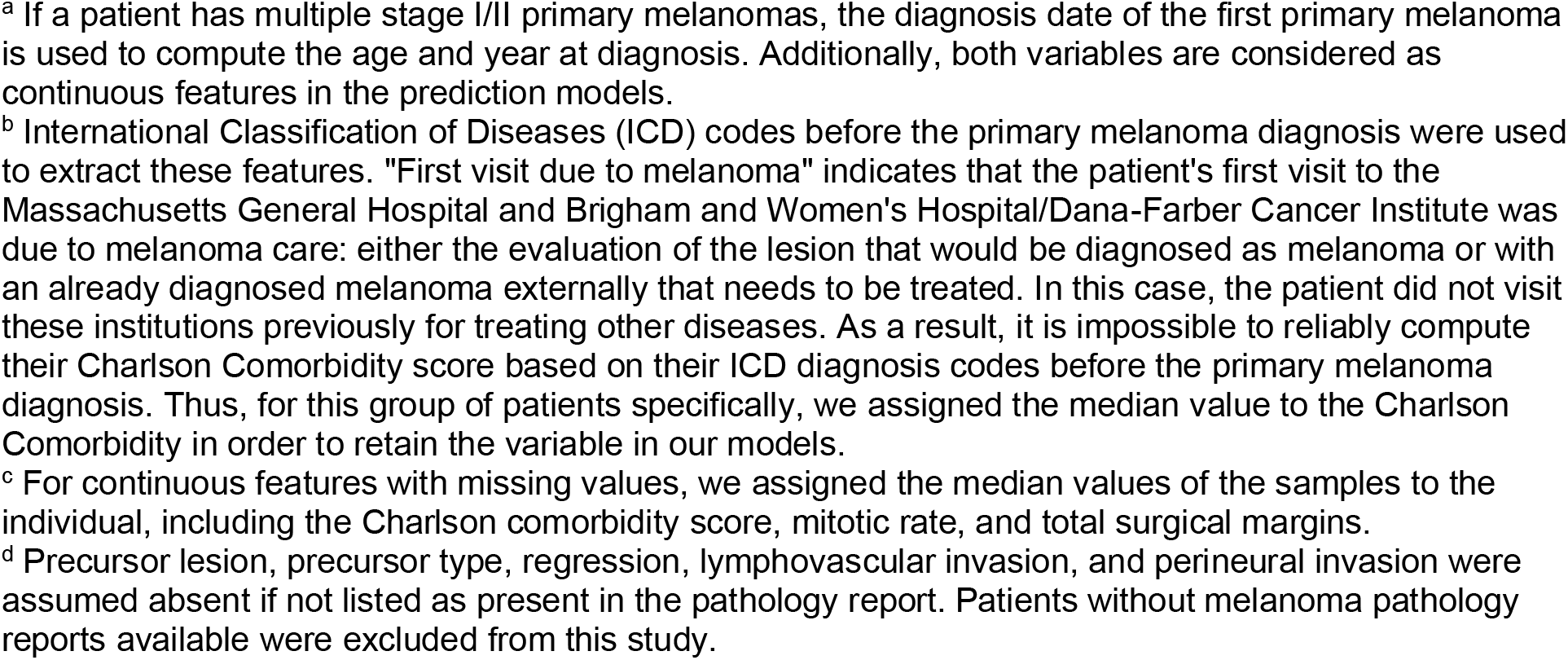
Characteristics of the study population for all extracted variables

**Supplementary Figure 1.**
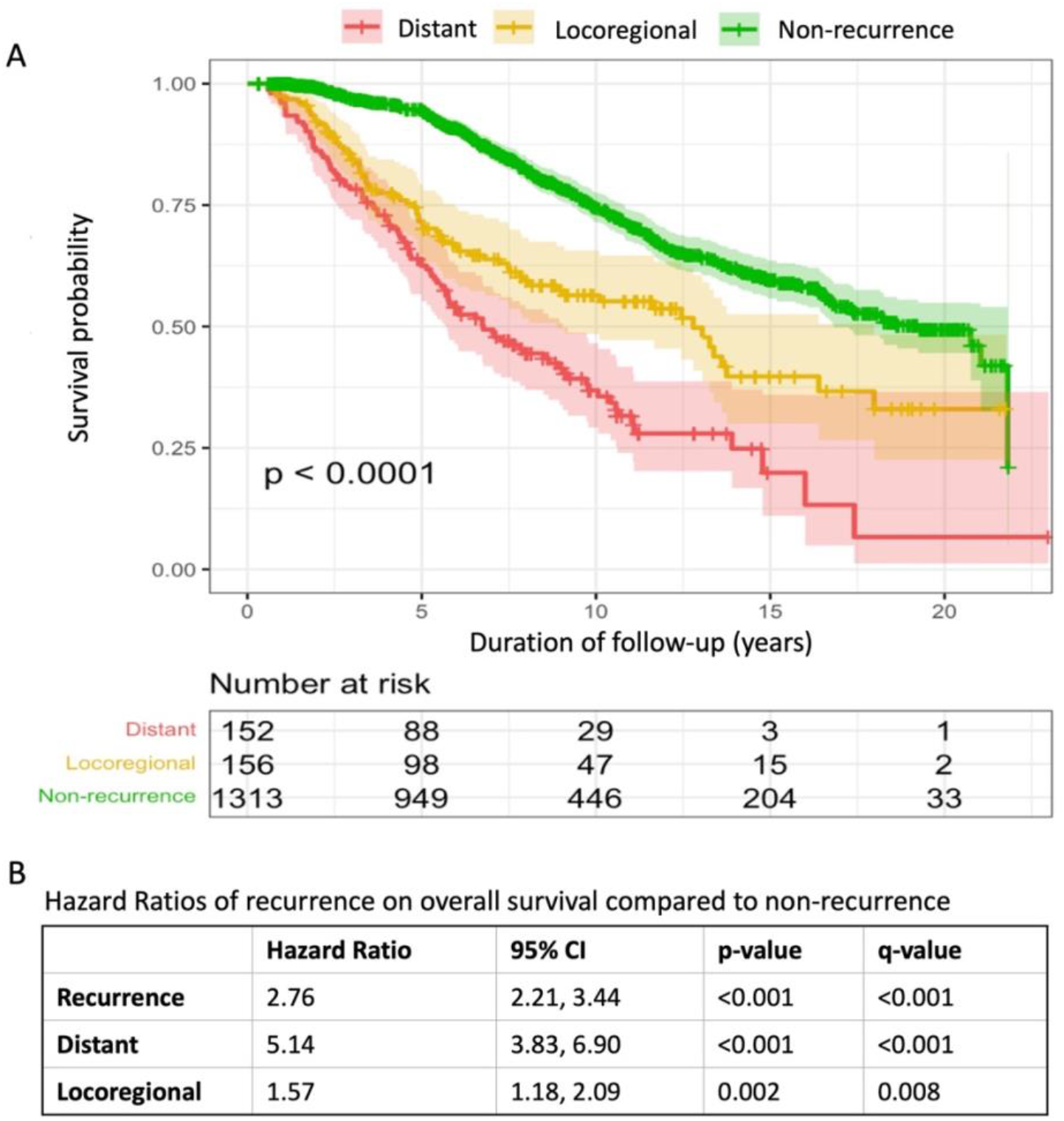
Association between recurrence-free survival and overall survival.

**Supplementary Figure 2.**
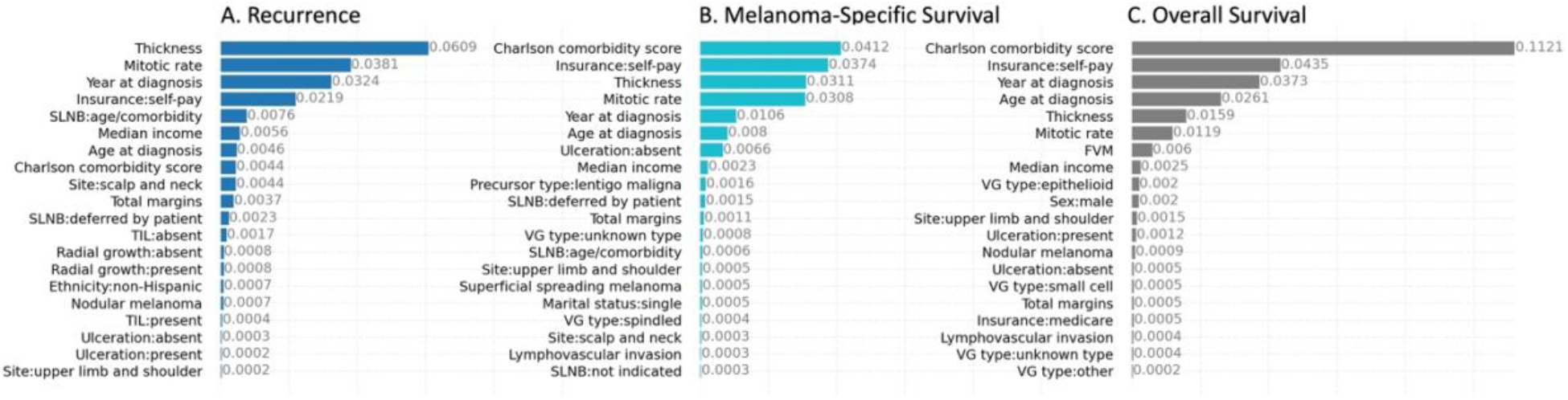
Feature importance in the melanoma recurrence and overall survival prediction models by GBS. SLNB: sentinel lymph node biopsy; TIL: tumor infiltrating lymphocytes; VG: vertical growth.

**Supplementary Figure 3.**
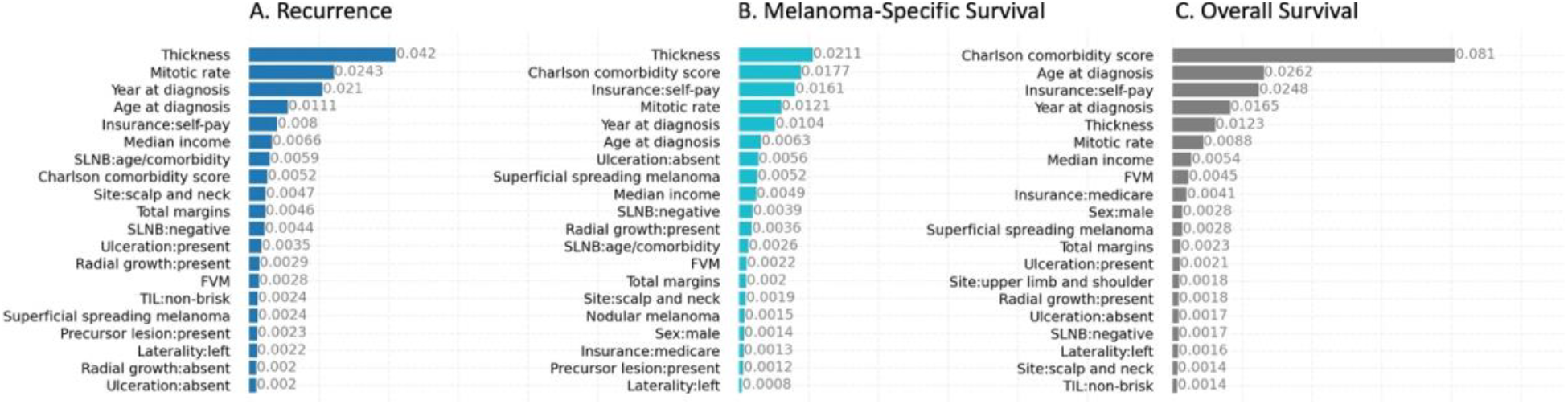
Feature importance in the melanoma recurrence and overall survival prediction models by RSF. SLNB: sentinel lymph node biopsy; TIL: tumor infiltrating lymphocytes; VG: vertical growth.

